# Video Consultation in Lung Transplant Recipients during the COVID-19 Pandemic

**DOI:** 10.1101/2020.05.12.20093799

**Authors:** Moritz Z. Kayser, Christina Valtin, Mark Greer, Bernd Karow, Jan Fuge, Jens Gottlieb

## Abstract

**Background:** The COVID-19 pandemic has disrupted health care systems worldwide. This is due to the demand for medical resources in other areas as well as concern for the risk of nosocomial SARS-CoV-2 exposure. The interruption of routine care is especially problematic for patients with chronic conditions requiring regular follow-up, such as lung transplant recipients. New methods like telemedicine are needed to provide care to these patients.

**Methods:** A retrospective analysis of video consultations (VC) in comparison to on-site visits (OSV) was performed during a six-week period in a lung transplant center in Germany. VC included a structured work-up questionnaire and vital sign documentation.

**Results:** During the 6-week study period, 75 VC were performed for 53 patients and 75 OSV by 51 patients occurred. By the end of our study period, 77% of physician-patient contacts occurred via VC. Overall, physician-patient consultations were reduced by 47% in comparison to an equivalent time frame in 2019. In 62% of cases, VC resulted in a concrete clinical decision. For two VC patients, the indication for inpatient admission was established during the consultation. One COVID-19 patient in home quarantine was admitted due to respiratory failure detected by VC. Patient satisfaction with VC was high.

**Conclusions:** By transitioning to VC, OSV for lung transplant patients during the COVID-19 pandemic was reduced. VC was well received by the majority of patients. This technology can be adopted to provide care for a wide range of chronic illnesses.

## Introduction

The emergence of the COVID-19 pandemic has disrupted health care systems worldwide^1^. Due to challenges posed by the virus itself as well as redistribution of health care resources during the pandemic, established procedures in outpatient care for those with chronic conditions have been interrupted^2-4^.

The American Medical Association (AMA) has published recommendations for the care of patients during the COVID-19 pandemic, including guidelines for remote consultation^5^. In the U.S., telemedicine has been successfully employed to provide care in a wide range of specialties^6^. Patients with chronic respiratory conditions like chronic obstructive pulmonary disease (COPD) ^7^ or interstitial lung diseases (ILD)^8^ are especially endangered by COVID-19^9,10^, and traveling to specialized health care facilities puts them at additional risk^11^. Lung transplant (LTx) recipients are extremely vulnerable due to their intense immunosuppressive therapy^12^. LTx patients require complex, continuous medical surveillance due to polypharmacy, co-morbidities, and the risk of allograft rejection^13-15^. Thus, follow-up care by a physician with expertise in lung transplantation cannot be deferred until after the pandemic, nor can it be completely replaced by home visits from general practitioners. This conundrum highlights the need for new strategies.

Video consultations (VC) have generated increased interest following the rise of video conference programs like Skype and Zoom in other sectors of the economy for professional and personal use^16^. However, use of conventional video conferencing tools is limited by data protection regulations due to concerns for patient privacy^17^. Recently, specialized video conference software that protect doctor-patient confidentiality have become more widely available, making VC an attractive option^18^. The object of this study is to analyze the clinical impact, technical feasibility and patient satisfaction of a rapidly-implemented online VC system as a partial replacement for OSV for LTx patients during the pandemic.

## Materials and Methods

A retrospective analysis was performed in a high volume LTx program during the COVID-19 pandemic in Germany. VC was conducted from calendar week 12 (beginning on March 16th, 2020) to calendar week 17 of 2020. Clinical decisions made based upon VC, technical feasibility, and patient satisfaction with this telemedicine program were investigated.

The reason underpinning each VC was categorized as either “routine surveillance” (replacing a scheduled surveillance visit in a stable patient), “follow-up” (assessment of recently initiated/altered therapies or discussion of recent test results) or “clinical indication” (new symptom requiring assessment). Similar categorization was performed for OSV. Prior to the COVID-19 pandemic, patients were seen for on-site surveillance consultations at regular intervals of 3 to 12 months, depending on the time elapsed since transplantation. Similar intervals were chosen during the pandemic for surveillance VC. The decision between VC and OSV was made based on clinical judgement, including whether bronchoscopy was indicated. VC was preferred wherever possible. Criteria such as age or distance between patient’s residence and study site were not taken into account when choosing between VC and OSV

All LTx patients in our program that were either scheduled for routine surveillance or required a consultation due to emergent clinical developments during the period between week 12 and week 17 of 2020 were eligible for this study.

VC was conducted using a video consultation tool (https://www.sprechstunde.online/, *Deutsche Arzt AG*, Essen, Germany) developed for use in an outpatient setting and certified according to the European General Data Protection Regulation (GDPR). Technical requirements were a computer or smart device with a camera, microphone and speakers, as well as an internet connection. Aside from the video conference function itself, we also used the text chat platform and a built in GDPR-approved data file exchange tool. Patients were contacted by phone to schedule an appointment for VC in advance. For first-time VC, the necessary equipment and technical steps were explained during this initial telephone conversation and a downloadable information sheet was provided (Supplement 1). All patients received coaching by our staff via telephone in advance of their appointments in addition to the written step-by-step guide. Technical guidance was estimated by our staff to take 10-15 minutes per patient for the initial VC, with reduced guidance necessary during subsequent VC.

Patients in our program are routinely equipped with a pulse oximeter and home spirometer (AM1/2, eResearch Technology, Philadelphia, USA) for measurement of Forced Expiratory Volume in 1 Second (FEV1) and Forced Vital Capacity (FVC)^19^ and are advised to record these values daily (Supplement 2). Each patient was contacted again on the day of the VC by our administrative staff to arrange an exact time for the consultation. They were then sent an invitation with a link to the VC online software by e-mail or text message. In addition, a standardized online symptom questionnaire was sent to patients (SoSciSurvey GmbH, Munich, Germany) to be filled out prior to the VC. VC was scheduled in 30-minute intervals with 10 minutes for staff to prepare between appointments. During the first part of each VC, the patient was asked about clinical condition, quality of life, changes in medication, and symptoms of infection. Additionally, daily vital sign documentation was examined (Table 1). Respiratory rate was measured using visual observation of the patient at rest by adjusting the camera to view the lower thorax. Breathing actions were counted via a tap counter app for one minute (https://apps.apple.com/at/app/tap-counter-with-sets/id1247135729, Philip Braham). Additionally, pulse rate and blood oxygen saturation (S_p_O_2_) were measured when available and home spirometry results were recorded. The second phase of the consultation focused on the patient’s current complaints, recent medical history and medication plan. Patients were invited to ask questions and a follow-up appointment (VC or OSV) was arranged. Examples of images taken during VC are shown in Figure 1 (**Not included in this pre-print manuscript in accordance with MeDxriv’s policy on patient images**).

**Table 1:** Patient assessment during video consultation. Summary of medical history and vital signs gathered systematically during VC

| Patient questionnaire | Vital signs |
| --- | --- |
| quality of life (visual analogue scale 0-10) | FEV1 home spirometry* |
| general state of health:<br>improved/stable/worsened | respiratory rate |
| coughing/sputum, yes/no, onset | pulse, regular/irregular |
| myalgia, malaise, yes/no, onset | blood pressure |
| runny or stuffy nose, yes/no, onset | oxygen saturation |
| sore throat, hoarseness, yes/no, onset | body temperature |
| fatigue, yes/no, onset | body weight |
| change in medication, yes/no, detail |  |
| physical fitness (flights of stairs without rest) |  |
| local lab report if available |  |
| current dose of calcineurin inhibitor |  |

\*FEV1- forced expiratory volume in one second

**Figure 1:** Sample photos of video consultation. **Not included in this pre-print manuscript in accordance with MeDxriv’s policy on patient images**.

After the VC, electronic documentation was completed in a clinical and scientific database (FileMaker Pro, Claris International Inc, Santa Clara, CA, USA). Following the interview, patients were sent an anonymous questionnaire via e-mail to ascertain their satisfaction with VC. Questionnaire data was exported from SoSciSurvey into SPSS Statistics data format. Statistical analysis was performed using IBM SPSS v26 (IBM SPSS Statistics, Armonk, NY, USA). Categorical variables are presented as numbers (n) or percentages (%), and continuous variables as median and interquartile ranges (IQR), unless indicated otherwise. Statistical significance between the groups was assessed using χ^2^ or Mann-Whitney U test as appropriate. A P-value of <0.05 was considered statistically significant. All tests were two sided. For demographic data, patients were sorted into groups depending on the type of first visit during the study period.

This study was performed in accordance with the ethical guidelines of the 1975 Declaration of Helsinki. All patients provided informed consent prior to transplantation allowing the use of their data for scientific purposes, as approved by the Ethics Committee of Hannover Medical School (Ethics Committee Vote Nr. 2923-2015). According to our Ethics Committee, additional approval was not necessary, as data acquisition was retrospective and observational, data was anonymized and the study relied on information collected as part of routine care. Patients seen in Figure 1 gave written consent for the publication of their images (**Not included in this pre-print manuscript in accordance with MeDxriv’s policy on patient images**).

## Results

During the 6-week study period, 75 VC were performed for 53 patients, compared to 75 OSV with 51 patients. The total number of consultations decreased by 47% during the COVID-19 crisis in 2020 compared to the equivalent time frame in 2019. The ratio of VC to OSV increased over the course of the pandemic (Figure 2). Fourteen patients had consultations by both VC and OSV during the study period. Patient characteristics are listed in Table 2. Both the median time post-transplantation and the median time between consultations were significantly shorter for on-site patients. The majority of OSV were required due to short-to-medium term post-transplant complications necessitating bronchoscopy. Average distance from patients’ home to our center was similar in both groups. Geographic distribution of patients is shown in Figure 3.

**Figure 2:**
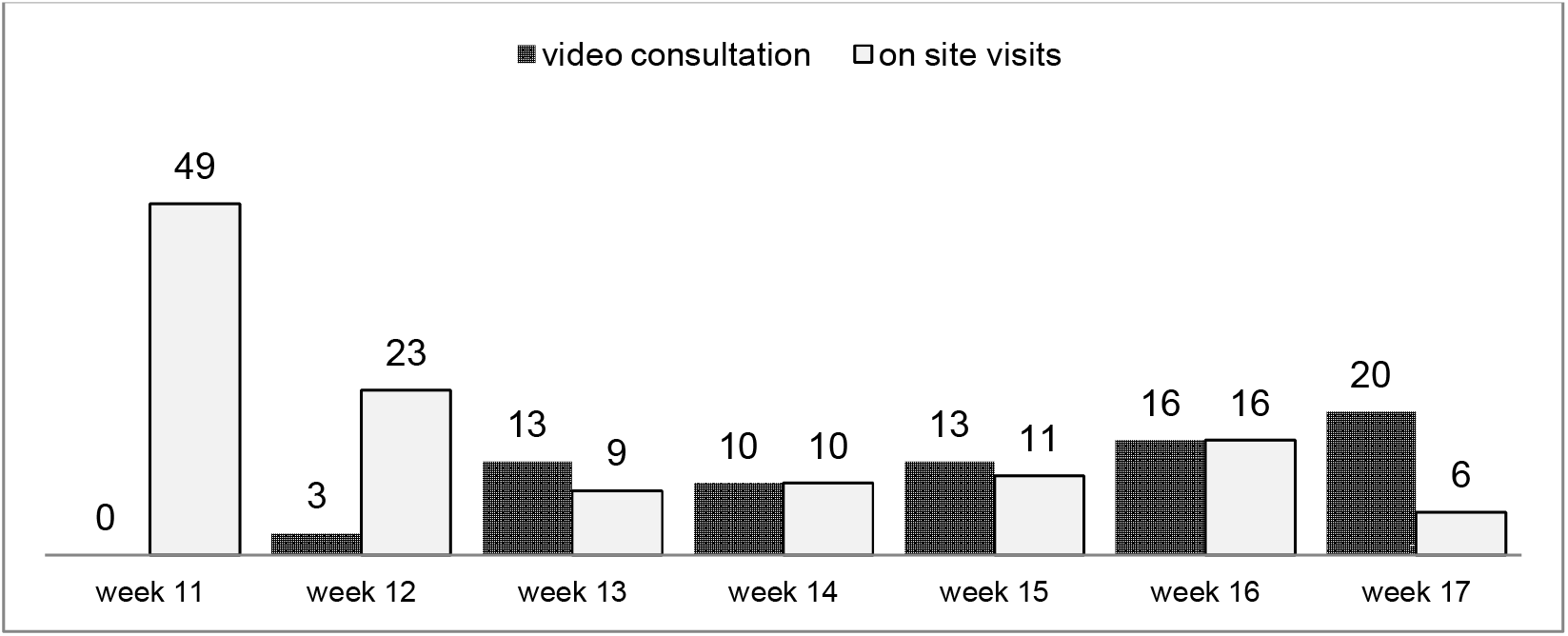
Number of physician-patient consultation per week during study period. Breakdown of VC and OSV by calendar week. Study period begins March 16^th^, 2020, corresponding to calendar week 12.

**Figure 3:**
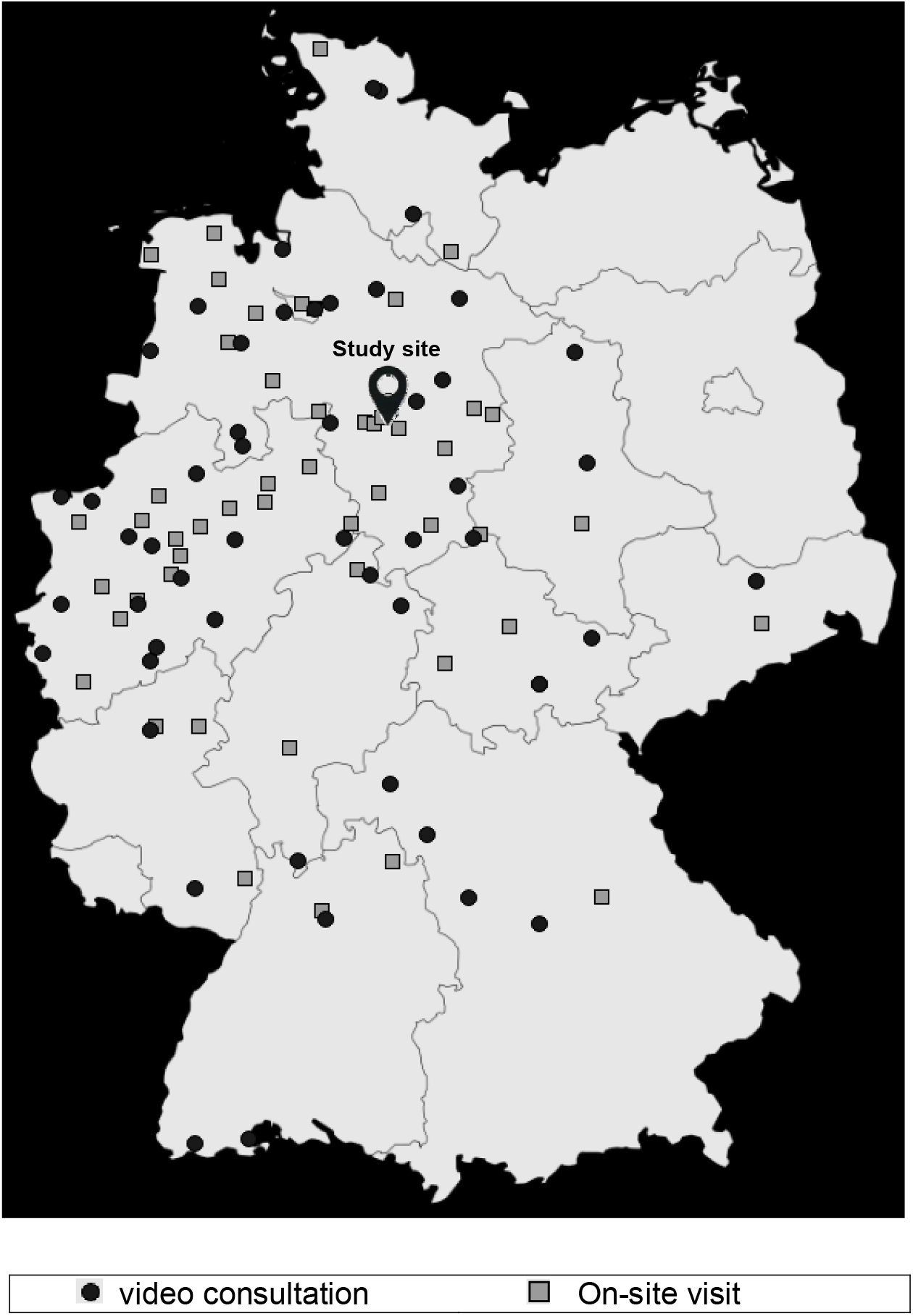
Geographic distribution of patients. Map depicting geographic distribution of patients’ location. Oval marker denotes study site (Hannover Medical School). Grey lines mark state borders.

**Table 2:**
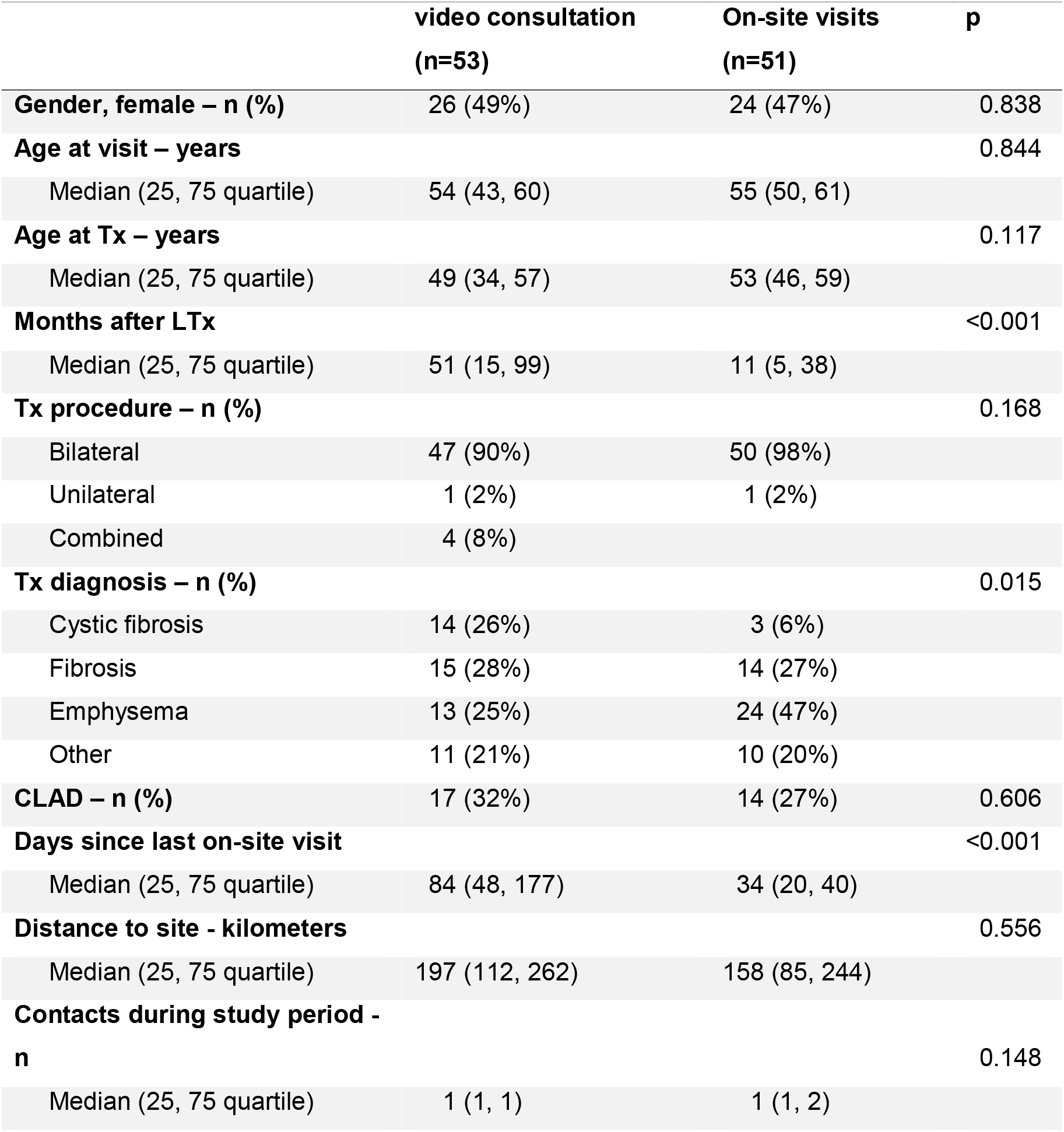
Patient demographics. LTx = Lung Transplantation, CLAD = Chronic Lung Allograft Dysfunction. P-values from comparisons between VC group and on-site visit group. Statistical significance between the groups was assessed using X^2^ for categoric variables or Mann-Whitney U test for metric variables. A P-value of <0.05 was considered statistically significant. All tests were two sided.

|  | video consultation<br>(n=53) | On-site visits<br>(n=51) | p |
| --- | --- | --- | --- |
| **Gender, female – n (%)** | 26 (49%) | 24 (47%) | 0.838 |
| **Age at visit – years** |  |  | 0.844 |
| Median (25, 75 quartile) | 54 (43, 60) | 55 (50, 61) |  |
| **Age at Tx – years** |  |  | 0.117 |
| Median (25, 75 quartile) | 49 (34, 57) | 53 (46, 59) |  |
| **Months after LTx** |  |  | <0.001 |
| Median (25, 75 quartile) | 51 (15, 99) | 11 (5, 38) |  |
| **Tx procedure – n (%)** |  |  | 0.168 |
| Bilateral | 47 (90%) | 50 (98%) |  |
| Unilateral | 1 (2%) | 1 (2%) |  |
| Combined | 4 (8%) |  |  |
| **Tx diagnosis – n (%)** |  |  | 0.015 |
| Cystic fibrosis | 14 (26%) | 3 (6%) |  |
| Fibrosis | 15 (28%) | 14 (27%) |  |
| Emphysema | 13 (25%) | 24 (47%) |  |
| Other | 11 (21%) | 10 (20%) |  |
| **CLAD – n (%)** | 17 (32%) | 14 (27%) | 0.606 |
| **Days since last on-site visit** |  |  | <0.001 |
| Median (25, 75 quartile) | 84 (48, 177) | 34 (20, 40) |  |
| **Distance to site - kilometers** |  |  | 0.556 |
| Median (25, 75 quartile) | 197 (112, 262) | 158 (85, 244) |  |
| **Contacts during study period -**<br>**n** |  |  | 0.148 |
| Median (25, 75 quartile) | 1 (1, 1) | 1 (1, 2) |  |

VC were distributed evenly between routine surveillance (24/75, 32%), follow-up (27/75, 36%) and clinical indication (24/75, 32%) (Table 3). Among OSV, 14/75 (19%) were for routine surveillance, 6/75 (8%) for follow-up, and 55/75 (81%) due to clinical indication. Most surveillance OSV (69%) took place during the first week of the study period. As time went on and our telemedicine program became more established, a decrease in routine surveillance visits conducted on-site was observed. For comparison, between calendar weeks 12 and 17 of 2019, 280 lung transplant recipients were seen on-site. Consultations scheduled due to clinical indication accounted for 19% of visits. The remaining 81% of visits were routine surveillance consultations. Of these 280 patients, 37% underwent bronchoscopy. Management of airway complications accounted for 33% of all bronchoscopies performed. VC due to clinical indication in 2020 were more frequent than indication-driven OSV during the historical control period in 2019.

**Table 3:**
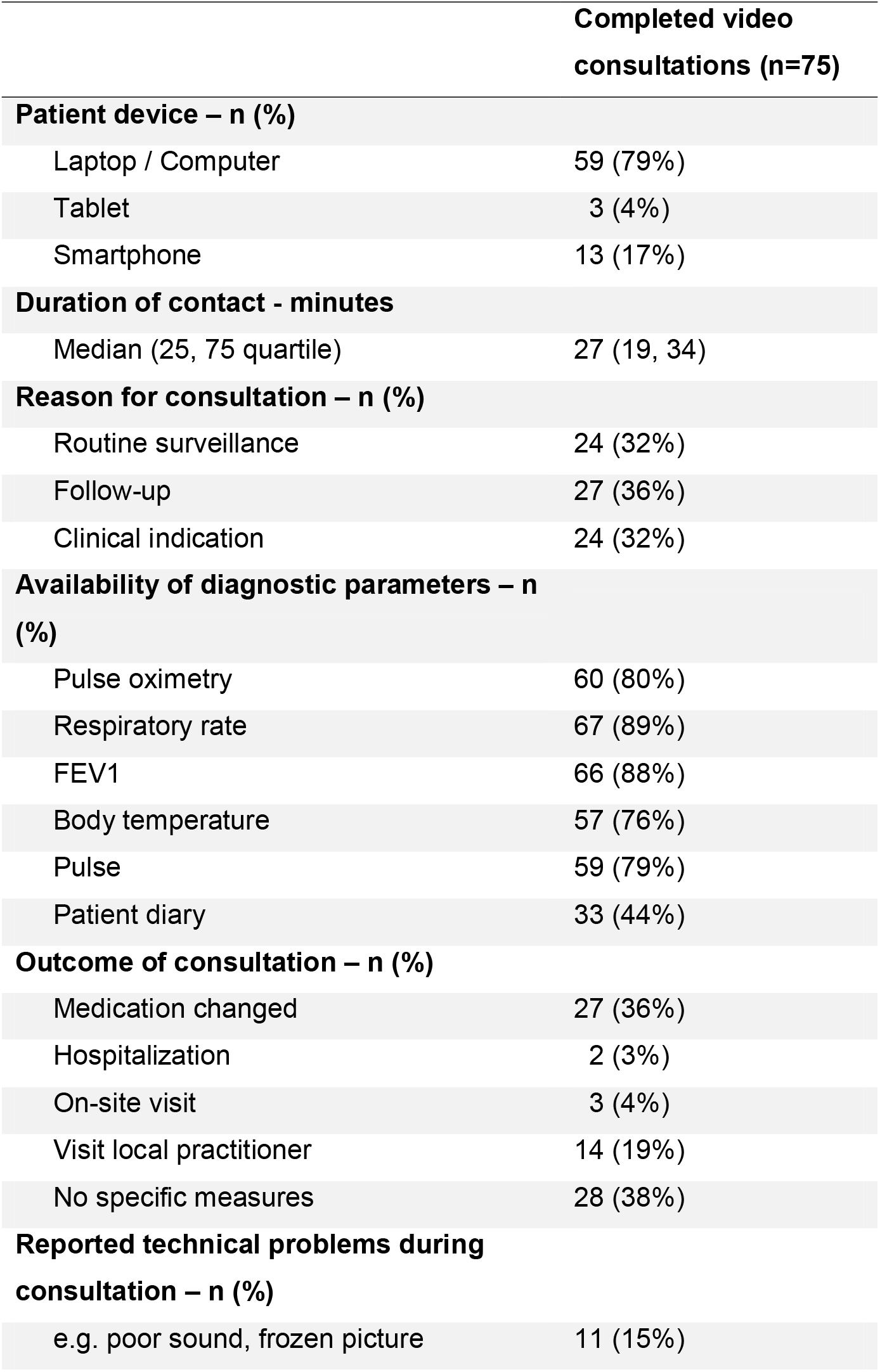
Summary of video consultations. Summary of video consultations. FEV1 = Forced expiratory one second capacity

|  | Completed video consultations (n=75) |
| --- | --- |
| **Patient device – n (%)** |  |
| Laptop / Computer | 59 (79%) |
| Tablet | 3 (4%) |
| Smartphone | 13 (17%) |
| **Duration of contact - minutes** |  |
| Median (25, 75 quartile) | 27 (19, 34) |
| **Reason for consultation – n (%)** |  |
| Routine surveillance | 24 (32%) |
| Follow-up | 27 (36%) |
| Clinical indication | 24 (32%) |
| **Availability of diagnostic parameters – n (%)** |  |
| Pulse oximetry | 60 (80%) |
| Respiratory rate | 67 (89%) |
| FEV1 | 66 (88%) |
| Body temperature | 57 (76%) |
| Pulse | 59 (79%) |
| Patient diary | 33 (44%) |
| **Outcome of consultation – n (%)** |  |
| Medication changed | 27 (36%) |
| Hospitalization | 2 (3%) |
| On-site visit | 3 (4%) |
| Visit local practitioner | 14 (19%) |
| No specific measures | 28 (38%) |
| **Reported technical problems during consultation – n (%)** |  |
| e.g. poor sound, frozen picture | 11 (15%) |

A minority of VC (38%) did not lead to specific action such as medication change. In 62% of cases, VC led to a concrete clinical recommendation. Fourteen consultations (19%) led to the recommendation for further diagnostic steps by the primary care physician such as laboratory testing or imaging studies. Twenty-seven consultations (36%) led to a change in medication. Three patients (4%) were brought in for an emergency OSV, due to findings on VC. In each of these three cases, acute allograft rejection was subsequently diagnosed. Two patients (3%) were admitted for inpatient care due to symptoms identified during VC (Table 2). Two VC occurred while patients were in home quarantine due to COVID-19. In one case, a 47-year-old male had previously presented to his general practitioner for cough and mild dyspnea and tested positive for SARS-CoV-2. We therefore arranged a VC during home quarantine. On VC, we observed respiratory distress and a low SpO^2^ of 81% on room air. Proper function of the pulse oximeter was checked during the VC by measuring the SpO^2^ of the patient’s father, who showed normal values. We arranged the immediate admission of this patient to his local hospital. His clinical condition worsened drastically within the next 48 hours, culminating in ICU admission, invasive ventilation and ECMO cannulation. In another case of VC leading to hospitalization, a 56-year-old female showed signs concerning for pleural empyema during a VC following a similar episode three weeks earlier that had been treated surgically. Due to findings on VC, this patient was admitted to our hospital for surgical revision the next day.

Of the 53 patients who received VC, 37 (70%) responded to a questionnaire on their satisfaction with the experience. Among the patients participating, the vast majority reported a high degree of satisfaction. The clinical aspects of the consultation were rated more favorably than the technical ones (Figure 4).

**Figure 4:**
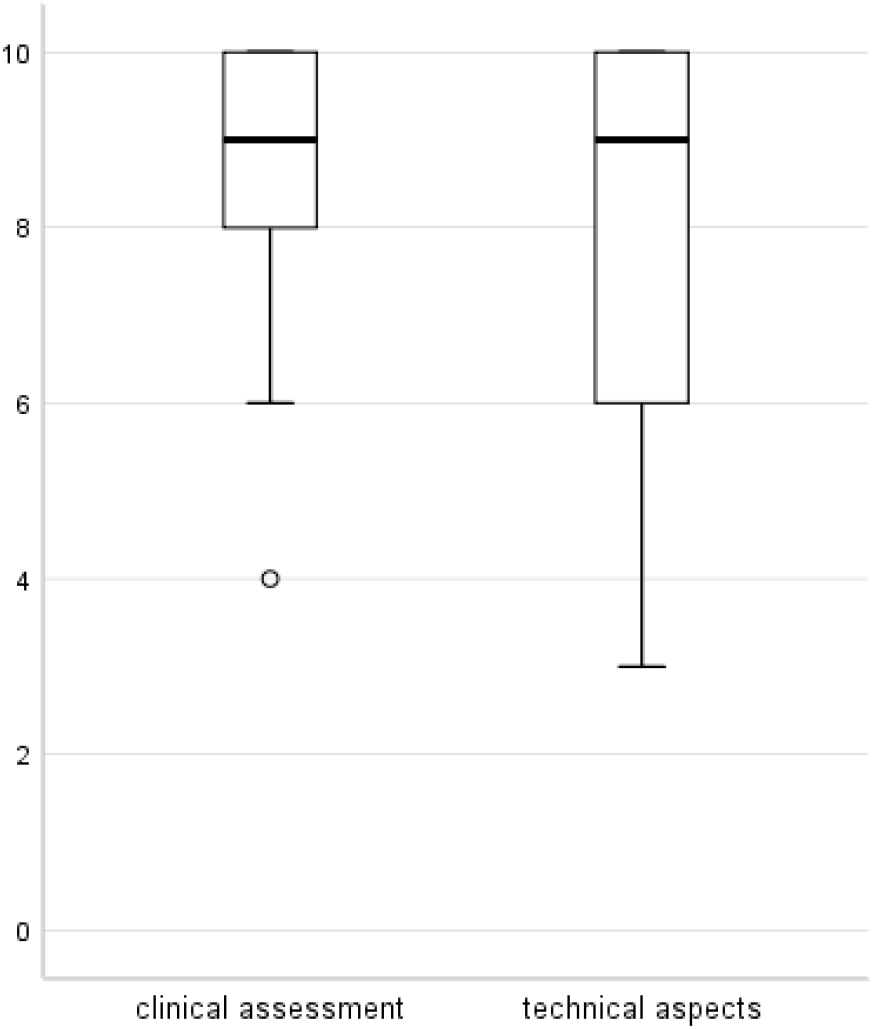
Patient satisfaction with video consultation. **Patient-reported satisfaction (n=38)** Boxplots show patient satisfaction as measured on a visual analogue scale (VAS)

In addition to our written guide, which outlines step-by-step procedures and includes a troubleshooting section, all of our patients received some level of assistance from our administrative coordinators via telephone in establishing VC. Minor technical issues on the patient’s end such as poor sound quality or intermittent screen freezing were reported during 15% of VC (Table 3). Initially, we also encountered in-hospital technical difficulties caused by the VC tool triggering our institution’s firewall, which required intervention by our IT staff. The high data volume required for VC also challenged our institution’s internet connection, especially during peak use times. Scheduling VC in the afternoon partially alleviated this problem.

In 8 patients, video consultation failed due to either patients’ lack of necessary equipment or due to technical difficulties. Three patients refused VC for personal reasons (Table 4). Altogether, VC was successfully established in 75/86 (87%) cases. The median age of patients where VC was not established was 62 years, 8 years older than the median age of patients with whom VC was successfully implemented.

**Tab. 4:** Summary of attempted contacts.

|  | Attempted video consultations (n=86) |
| --- | --- |
| Completed video consultations – n (%) | 75 (87%) |
| Failed video consultations – n (%) | 11 (13%) |
| Reason for failure – n (%) |  |
| Technical problems | 6 (55%) |
| Missing devices | 2 (18%) |
| Patient refused video consultation | 3 (27%) |

## Discussion

To our knowledge, this is the largest study on the implementation of VC in respiratory medicine during the COVID-19 pandemic. VC increased in our center for follow-up care post LTx during the study period while OSV declined. One third of VC occurred due to clinical indication. Overall satisfaction with VC was high.

There is currently one published study evaluating VC in transplant patients. In a randomized controlled trial, the readmission rate in 100 recent liver transplant recipients was lower in patients receiving telemedicine in addition to standard of care (SOC) compared to patients receiving SOC only^20^.

Outside the field of transplant medicine, VC has been studied in a variety of clinical contexts. In the home management of 35 newborns with severe heart defects, VC proved superior to both SOC and telephone consultation in reducing the rate of readmission. This study reported technical difficulties in 17% of VC, which is similar to the 15% rate we observed^21^. A majority of studies comparing VC to OSV have been conducted in the field of mental health^22^. While conclusions from these studies are limited by small sample size, the reported patient satisfaction rates are high^23^.

Recently, an American group reported on the implementation of VC in the care of 38 cystic fibrosis patients during a 4-week interval during the COVID-19 pandemic^24^. This study employed a commercial video chat program that does not conform to European Union GDPR requirements. In this study, VC was performed exclusively in patients without symptoms of infection, leading to 2 changes in medication and 1 OSV as a consequence of VC. Patient satisfaction was not assessed.

During the COVID-19 pandemic, a rapid transition to telemedicine was necessary to guarantee the continued care of our LTx patients while limiting their potential exposure to SARS-CoV-2. The observed drop in patient appointments, virtual or in person, during the pandemic reflects an understandable lag in implementing telemedicine rapidly and with little preparation. Until the beginning of the study period described here, none of our patients were seen via VC and no infrastructure for this transition was in place. The precipitous reduction in OSV seen in week 11 and 12 and the subsequent steady rise in VC speaks to the workability of a swift transition (Figure 2). However, specific patient populations continue to require care that cannot be delivered by VC. For example, graft hypoperfusion in the early stage post LTx can lead to the development of airway complications like fibrinous bronchitis and bronchial stenosis, requiring repeated interventional bronchoscopy^25^. Likewise, the diagnosis of acute allograft rejection often requires bronchoscopic lung tissue biopsy^14^. Among clinically indicated OSV during the study period, the vast majority were for such endoscopic procedures.

The success of VC was facilitated by a system of patient-owned monitoring devices (home spirometry, pulse oximetry) to objectively assess patients’ vital signs. Thus, we were able to identify hypoxemia in a COVID-19 patient and initiate hospital admission before severe clinical deterioration occurred. The implementation of a standardized symptom questionnaire helped lay the groundwork for VC. The measurement of respiratory rate by VC proved a valuable tool in the clinical assessment of patients. This highlights the advantages of VC in fields that rely heavily on the visual assessment of patients, such as respiratory medicine and infectious disease.

The majority of VC resulted in either a change of medication or a recommendation for further diagnostic testing, which required our center’s expertise due to the complexity of this patient population. We believe this approach can be translated to other chronic respiratory diseases requiring specialized medical care. However, a close collaboration with physicians local to the patient’s place of residence remains paramount, especially when further diagnostic steps or procedures are required. In the context of the pandemic making travel difficult, as well as the risk of nosocomial SARS-CoV-2 infection during a visit to a health care facility, VC represents a balance between access to specialist expertise and exposure prevention.

The vast majority of patients were satisfied by both the clinical the technical aspects of VC. This high rate of satisfaction is likely due to the structured approach we employed to ensure a smooth logistical and technical process. However, the lack of response to patient satisfaction questionnaires in 30% of VC patients may have led to an overestimation of patient satisfaction with VC. Reasons for non-response to the patient questionnaire may include disinterest or technological fatigue. One out of eight attempted VC was unsuccessful due to patients’ lack of equipment, technical difficulties, or patient refusal. Patients with whom VC was not established tended to be older, underlining the concern that patient age poses a relevant barrier to telemedicine^26^.

It is important to note that Germany has poor digital infrastructure compared to other western European countries, particularly in the health care sector^27^. However, our overall success in implementing VC highlights the willingness of our patients to transition to a telemedicine approach. VC may meet with even greater success in countries with a more advanced digital infrastructure. Direct data transmission from patient-owned monitoring devices, as is already common in a number of commercial health and fitness devices, could facilitate follow-up care in the future^28^. There are a variety of additional tools in development to allow more diagnostic steps to be taken remotely. For example, a Belgian group is currently developing a system of remote pulmonary auscultation^29^. In time, some combination of OSV and VC will likely become standard in a wide range of medical specialties. The current COVID-19 crisis and the attendant shift to telemedicine may well catalyze a new level of acceptance of this technology by patients and physicians alike.

In conclusion, we demonstrate that VC can be a valuable tool in the effort to provide quality health care to LTx patients during the COVID-19 pandemic. Our center’s experience and the roadmap outlined above provide a framework for the implementation of telemedicine for patients with chronic respiratory diseases. With a possible SARS-CoV-2 vaccine still years away, the risk of morbidity and mortality from COVID-19 will inform decision making in health care systems worldwide for the foreseeable future. In this context, VC will play an important role for physicians treating chronically ill patients, and research investigating best practices and new models of care in this area are more relevant than ever.

## Data Availability

There are no supplementary data sets pertaining to this manuscript online.
Our video consultation guide (Supplement 1) and vital parameter log-sheet (Supplement 2) are attached below (German).

https://www.mhh.de/fileadmin/mhh/pneumologie/downloads/Videosprechstunde_Anleitung.pdf

https://www.mhh.de/fileadmin/mhh/pneumologie/downloads/pdf/mhh_ltx_tagebuch.pdf

## Acknowledgements

We would like to thank our administrative coordinators Konstantina Zang-Pappa, Linda Häsler, Anita Fuhrmann and Bianca Metzdorf for their patience with both patients and physicians. Their persistence and efficiency made our clinic’s transition to telemedicine possible.

**Figures and Tables: Video Consultation in Lung Transplant Recipients during the COVID-19 Pandemic**

